# Operationalising the WHO call for integrated emergency care through a national ambulance alliance network: implementation experience and lessons from Addis Ababa, Ethiopia (PRECOS-1)

**DOI:** 10.64898/2026.07.22.26358728

**Authors:** Peniel K. Dula, Katherine R. Iverson, Aklilu Azazh, Biniyam Kifleyohanes, Emnet T. Shimber, Ermiyas D. Asefa, Habtamu Mengie, Natnael Zenebe, Oumer A. Nuru, Rahel A. Assefa, Tenbite Daniel, Woldesenbet Waganew, Tewodros Kifleyohanes, Elubabor Buno Teko, Kokeb D. Belihu, Yared Boru, Fitsum K. Belachew, the Ethiopian Ambulance Alliance Network (EAAN) Collaborative

## Abstract

Prehospital emergency care in many African cities is constrained not by ambulance scarcity but by fragmentation: multiple uncoordinated provider types operating parallel dispatch systems, with no shared data and no capacity to measure system performance. Despite the World Health Assembly’s 2023 resolution on integrated emergency care (WHA 76.2) and Ethiopia’s Health Sector Transformation Plan II (HSTP-II), prehospital coordination has remained a critical missing link. In response, Ethiopia developed the Hospital Emergency Assistance and Response Tracking System (HEARTS) under the Ethiopian Ambulance Alliance Network (EAAN), a governed, multi-provider federation established through consultation with the Network for Perioperative and Critical Care (N4PCc), Federal Ministry of Health, the Addis Ababa city administration fire and disaster response team, a humanitarian NGO provider, and dedicated private ambulance providers. HEARTS provides real-time fleet tracking, unified call processing, community access via a dedicated mobile application, and performance dashboards disaggregated by sub-city and provider type. This paper reports the first phase of the Prehospital Care Outcome Study (PRECOS-1), a programmatic research platform established to generate longitudinal evidence on prehospital coordination in Ethiopia and to inform replication across Africa. During the first operational phase in Addis Ababa (December 2025 to June 2026), HEARTS coordinated 1,403 trips across four provider types and 59 response units. The majority of trips were high- or critical-acuity (63.3%), with maternal and obstetric presentations forming the largest clinical category (44.3%). Median response time was 14.5 minutes (IQR 5.8 to 27.0; 90th percentile 75.0 minutes), the first unified prehospital performance baseline for this city. These findings demonstrate that a coordination-first approach to prehospital system strengthening is feasible in a fragmented low- and middle-income country setting, generate the measurement infrastructure for future improvement studies (PRECOS-2 onward), and offer a replicable model for African cities pursuing integrated emergency care.

## Introduction

Each year, an estimated 54 million deaths are attributable to conditions amenable to emergency care, with low- and middle-income countries (LMICs) bearing the greatest burden; more than 50% of these deaths may be preventable with quality emergency care systems (1,2). Road traffic injuries alone cause 10% of global mortality, representing the leading cause of death in young people aged 5 to 29 years, with the highest toll in LMICs (3). In 2023, the World Health Assembly responded with resolution 76.2 (WHA 76.2), renewing the global commitment to integrated emergency, critical, and operative care and calling for strengthened communication, transportation, and referral linkages across health systems (4). The WHO subsequently published operational guidance for prehospital ambulance systems, providing frameworks for governance, standardised documentation, and quality monitoring (5). Despite these commitments, most African cities face a structural paradox: the primary constraint on access to emergency care is not ambulance scarcity but fragmentation.

Across sub-Saharan Africa, fewer than 9% of the population is served by any emergency medical services (EMS) system (6). A 2023 systematic review of prehospital care in LMICs found that systems are overwhelmingly fragmented and uncoordinated, with most of the 87 reviewed studies reporting the absence of any structured service (7). A prior systematic review across 13 LMICs identified inadequate funding and a lack of coordination among existing services as the two most common barriers to prehospital development (8). In cities across Africa, multiple provider types typically operate in parallel: public agencies at federal, regional, and city-administrative levels; NGO humanitarian networks; private dedicated ambulance services; and hospital-based transport. Each maintains its own dispatch system, emergency access number, and paper records. The critical consequence is not merely inefficiency but invisibility: when providers share no data, system performance cannot be measured, and what cannot be measured cannot be improved. The WHO Emergency Care Systems Framework identifies this as a fundamental gap, noting that cross-cutting functions, including communication, data management, and quality monitoring, must link scene, transport, and facility care (9,10).

The potential of digital coordination platforms to address this gap is evidenced by experience elsewhere in the region. In Kenya, a multi-provider digital dispatch system integrated over 800 ambulance providers, substantially reducing response times and coordinating more than 47,000 calls by 2025 (11). In Rwanda, qualitative research documented that dispatchers’ inability to locate emergencies and limited inter-provider coordination were the primary barriers to EMS quality, precisely the functions that shared digital platforms address (12,13). The Global Prehospital Consortium, following a nine-round modified Delphi study, placed communication and coordination among the seven highest priorities for LMIC EMS development (14). Community willingness to call a formal ambulance rather than resort to informal transport is also recognised as a critical demand-side factor that coordination infrastructure directly influences (7,8,14).

Ethiopia presents the fragmentation paradox in stark form. With more than 4,000 public-sector ambulances (15). The country is not short of vehicles by population benchmarks, yet fewer than 10% of severely injured patients reach the hospital by ambulance (16) . In Addis Ababa, the capital, EMS use is low despite the availability of ambulances: only 20.3% of emergency patients arrive by ambulance, with most transported by taxi or private car (17). Long wait times deter use, and knowledge of emergency numbers significantly influences ambulance utilisation (18). Ethiopia’s HSTP-II (2020/21 to 2024/25) commits to improving access to emergency care (15),yet prehospital coordination remains a documented gap. A companion mixed-methods needs assessment (June to July 2025; 65 provider surveys at 93% response rate; 13 stakeholder interviews) documented system fragmentation across six domains: resource constraints, workforce limitations, dispatch and communication inefficiencies, inter-provider coordination failures, limited public awareness, and digital readiness barriers (19). Stakeholders described duplicate responses to some calls, while others went unanswered; private ambulances were turned away from public facilities; and there was a complete absence of system-wide performance data (19).

The Hospital Emergency Assistance and Response Tracking System (HEARTS) was developed to address this coordination gap at the city level, operationalising the integration mandate of WHA 76.2 and contributing to HSTP-II objectives (20). Built through iterative consultation with the Federal Ministry of Health, the Addis Ababa city administration, an NGO humanitarian provider, and private dedicated ambulance providers, HEARTS is the founding digital infrastructure of the Ethiopian Ambulance Alliance Network (EAAN), a governed federation of existing providers designed to render Ethiopia’s fragmented prehospital system integrated, measurable, and continuously improvable (Figure 1). The EAAN model follows the two-tier LMIC EMS framework endorsed by the African Federation for Emergency Medicine and recently reaffirmed by Sun, de Vries, and Mould-Millman: Tier-1 community-first responder care complemented by Tier-2 formal EMS, linked through a coordination infrastructure (21,22). The aim of this study is to describe the implementation of HEARTS and EAAN, present early operational findings from the first phase of the Prehospital Care Outcome Study (PRECOS-1), and outline lessons to inform replication in other fragmented prehospital systems.

**Figure 1:**
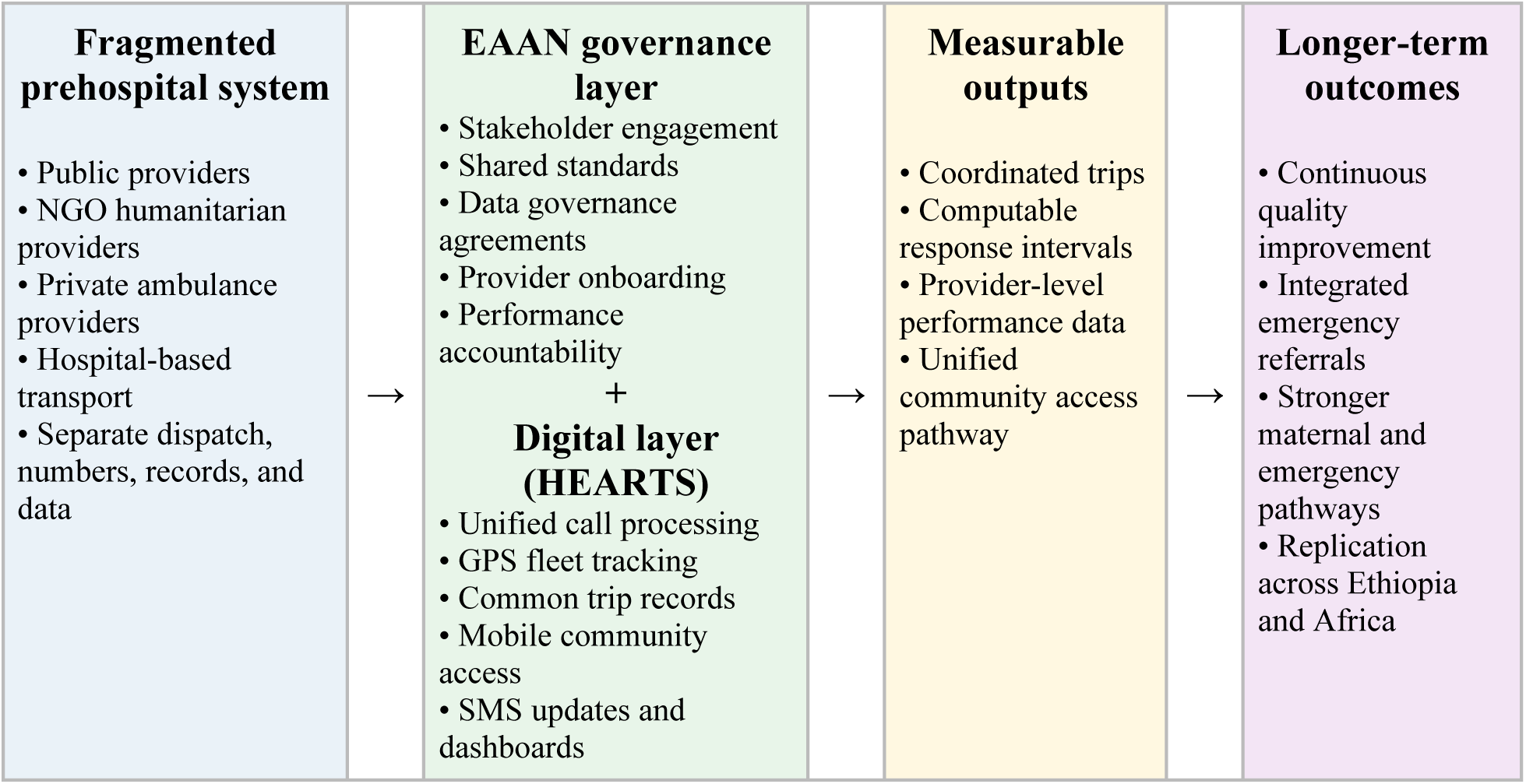
Conceptual framework for transforming fragmented prehospital ambulance services into an integrated, measurable emergency care system through EAAN and HEARTS.

## Methods

### Study design

PRECOS-1 is a mixed implementation and observational study combining three data sources: a pre-implementation mixed-methods needs assessment conducted between June and July 2025; a narrative account of HEARTS platform development, governance structure, and stakeholder engagement process; and a cross-sectional descriptive analysis of routine HEARTS platform operational data from the first active implementation phase. Reporting follows elements of the STROBE checklist for observational studies and the SQUIRE guidelines for quality improvement study reporting. The study is designated as the first phase of the PRECOS programmatic research platform.

### Setting

Addis Ababa, the capital of Ethiopia (estimated population of six million in 2025; 11 sub-cities) (23), Historically, five distinct prehospital provider layers operated independently: (1) federal and regional public services under the Ministry of Health; (2) the Addis Ababa Fire and Disaster Risk Management Commission (AAFDRMC), the principal city-administrative provider, operating sub-city dispatch stations across all 11 sub-cities; (3) a nationwide NGO humanitarian providers operating a volunteer ambulance networks; (4) private dedicated ambulance providers; and (5) hospital-based transport services. Each provider maintained its own dispatch centre, emergency access number, and paper-based patient care records, with no cross-provider data sharing or joint operational oversight.

### EAAN governance and HEARTS development

EAAN was established as a multi-stakeholder governing body under which each provider retains operational identity and ownership while committing to shared dispatch coordination, interoperability standards, and performance reporting. This governance model was designed following the clinician- and institution-led collaborative approach adopted by the NaPQIN peri-operative quality registry, led and coordinated by the same network (24). EAAN governance development proceeded through five sequential phases between 2023 and 2025: (1) landscape analysis and provider mapping across all 11 Addis Ababa sub-cities; (2) bilateral consultations with each provider type to document operational workflows, call volumes, and data systems; (3) co-design workshops to establish shared governance principles, data standards, and interoperability requirements; (4) formal regulatory endorsement and Ministry of Health consultation under the HSTP-II framework;(15) and (5) phased provider onboarding with training, technical support, and defined escalation pathways.

HEARTS was developed as EAAN’s digital infrastructure. The platform provides: (1) a unified emergency call reception and dispatch interface accessible via desktop and mobile application, including a patient-facing mobile application with real-time SMS status updates at each dispatch stage; (2) real-time GPS fleet tracking across all registered provider vehicles; (3) a common trip record capturing timestamps, incident type, acuity triage, clinical fields, and destination; and (4) real-time performance dashboards disaggregated by sub-city and provider type, displaying response time distributions, call volume trends, case mix, and provider performance comparisons. Alongside PRECOS-1, a modified Delphi consensus process is underway among prehospital emergency care experts in Ethiopia to refine and standardise the core outcome variables captured by the HEARTS platform, following the precedent of the Global Prehospital Consortium Delphi study on LMIC EMS priorities (14). Results of this process will be reported separately and will define the variable framework for PRECOS-2 onward. Future work will explore collaboration with EMS networks in the wider African region to extend this variable standardisation.

Community engagement was embedded in HEARTS implementation from the outset. A dedicated mobile application enables community members to request ambulances directly, track the responding vehicle in real time, receive automated SMS status updates and rate the services they received. Community outreach campaigns were conducted across Addis Ababa sub-cities to promote awareness of the unified emergency access point, with targeted emphasis on maternal and obstetric emergencies.

### Data source and study period

Routine operational data were extracted from the HEARTS platform as a de-identified export covering trips created between 25 December 2025 and 2 June 2026. Extracted variables included: provider organisation, response unit identifier, trip creation date and time, dispatch time, en route time, scene arrival time, scene departure time, and destination arrival time. Clinical variables included incident type, incident severity (Low/Medium/High/Critical), age, sex, and nature of case.

A data migration period (February 2026) was identified and excluded from the primary analysis on the basis of anomalous trip volume (1,799 records versus 52 to 529 in all other months) and markedly lower timestamp completeness (12% versus 63 to 88% in adjacent months). The analytic dataset comprised trips from all remaining months in the extraction period, yielding 1,403 trips over approximately 130 active operational days. Provider organisations are described by type to protect commercial confidentiality; the city-administrative public provider (AAFDRMC) is named as a government body, a convention standard in public health research.

Implementation challenges and organisational responses documented in the Results were derived from a fourth data source: prospective implementation records maintained by the EAAN team throughout the platform development and operational period (2023 to 2026). These records comprised field notes and observation logs captured during provider engagement activities, site visits, and training sessions across all 11 sub-cities; implementation monitoring records tracking provider onboarding status, platform activation, and escalated technical issues; and team meeting minutes and stakeholder correspondence documenting decisions made in response to emerging barriers. Findings from these sources were cross-referenced with patterns observed in the quantitative operational data, including documentation completeness rates and provider participation levels, to corroborate and contextualise the challenges reported.

### Statistical analysis

All analyses were descriptive. Categorical variables are reported as absolute frequencies and proportions. Continuous response interval variables are reported as medians with interquartile ranges (IQRs) and 90th percentiles, given their non-normal distributions confirmed by visual inspection of histograms. Response intervals were computed as: activation time (dispatch timestamp to en route timestamp); response time (dispatch timestamp to scene arrival timestamp); on-scene time (scene arrival to scene departure); transport time (scene departure to destination arrival); and total task time (dispatch to destination arrival). Intervals were excluded from the analysis if the computed value was negative or exceeded 720 minutes, as these were considered implausible and indicative of data entry errors. Provider-stratified analyses examined response interval distributions and acuity profiles by organisation type. Incident types were grouped into clinical categories, maternal and obstetric, cardiac, stroke or CVA, respiratory, trauma and accident, and other medical, based on the standardised classification of free-text entries in the HEARTS platform. Data completeness is reported for each variable as the proportion of records with a non-missing, valid entry. No imputation was performed for missing values. Analyses were conducted in Python 3.12 (Python Software Foundation).

### Ethics

The study was approved by the AHRI/ALERT Ethics Review Committee (protocol PO-039/25, ’Strengthening Prehospital Emergency Care in Ethiopia with HEARTS: A Hospital Emergency Assistance and Response Tracking System’, approval period 18 July 2025 to 17 July 2026). Patient-identifiable fields present in the raw platform export (name, date of birth) were excluded from all analyses.

## Results

### Provider integration

During the study period, HEARTS coordinated trips from four provider types across 59 response units (Table 1). AAFDRMC contributed 1,109 trips (79.0%), of which 77% were classified as high or critical acuity. A hospital-based provider contributed 232 trips (16.5%), of which 9% were high-or critical-acuity. A central coordination dispatch category accounted for 57 trips (4.1%), with 12% at high or critical acuity. Private dedicated ambulance providers collectively contributed 5 trips (0.4%), with 20% of those trips being high- or critical-acuity. The NGO humanitarian provider was not yet contributing trip data at the time of the data extraction. Across all providers, 63.3% of trips were high- or critical-acuity.

**Table 1.**
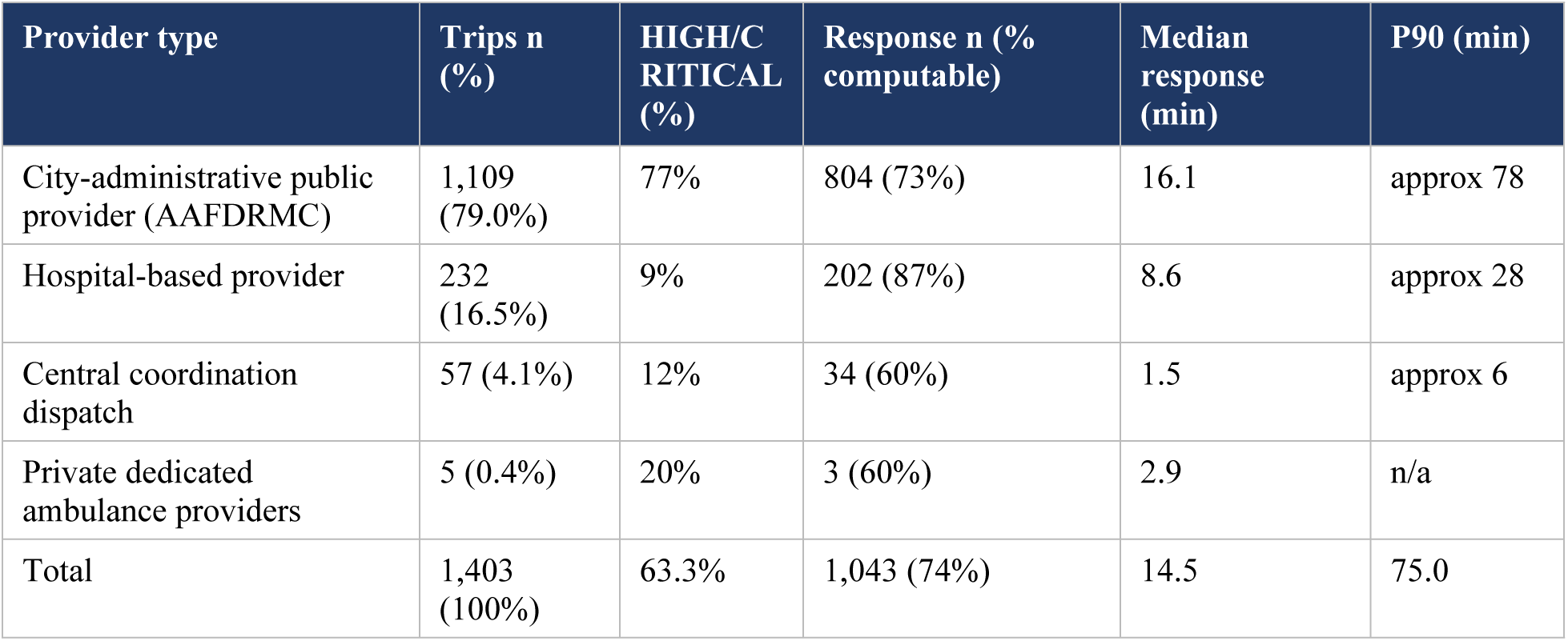
HEARTS operational profile by provider type, Addis Ababa, December 2025 to June 2026.

### Call volume and trip outcomes

Monthly call volume was 52 trips in December 2025 (partial month), 328 in January 2026, 529 in March 2026, 268 in April 2026, 210 in May 2026, and 16 in June 2026 (partial month). Of 1,403 trips, 1,089 (77.6%) were completed, with destination arrival recorded; 256 (18.2%) were cancelled; and 58 (4.1%) remained unresolved at the time of data extraction. Among cancelled trips, recorded reasons included: no ambulance available, no answer from the provider, and the caller used alternative transport.

### Clinical case mix

Maternal and obstetric presentations formed the largest clinical category, comprising 621 trips (44.3%). Other medical presentations accounted for 443 trips (31.6%), trauma and accident for 158 trips (11.3%), cardiac presentations for 73 trips (5.2%), respiratory presentations for 45 trips (3.2%), other or unknown for 43 trips (3.1%), and stroke or CVA for 20 trips (1.4%). Median estimated trip distance was 11.1 km (IQR 6.4 to 17.2). The detailed clinical case mix is presented in Table 2 below.

**Table 2.** Clinical case mix, HEARTS first operational phase, Addis Ababa.

| Clinical category | Trips n (%) |
| --- | --- |
| Maternal and obstetric | 621 (44.3%) |
| Other medical | 443 (31.6%) |
| Trauma and accident | 158 (11.3%) |
| Cardiac | 73 (5.2%) |
| Respiratory | 45 (3.2%) |
| Other or unknown | 43 (3.1%) |
| Stroke or CVA | 20 (1.4%) |
| Total | 1,403 (100%) |

A patient account from the HEARTS operational period, shared with full written consent, is presented below.

> *When I called the government ambulance service, they picked up immediately. The ambulance arrived in under five minutes. The SMS updates we received through HEARTS at each stage made the whole process clear and reassuring. It gave us confidence at a moment of stress. Because of this experience, I have been encouraging my friends and community to use ambulance services whenever they face emergencies instead of relying on informal transport.*

*Patient X (husband of patient X) shared with the full written consent of the patient and family*.

### Response intervals

Response intervals were computable for 1,043 trips (74.3%) for the response time measure. Timestamp completeness varied by interval type and by provider. Table 3 presents the full distribution of response intervals across all computable trips.

**Table 3.**
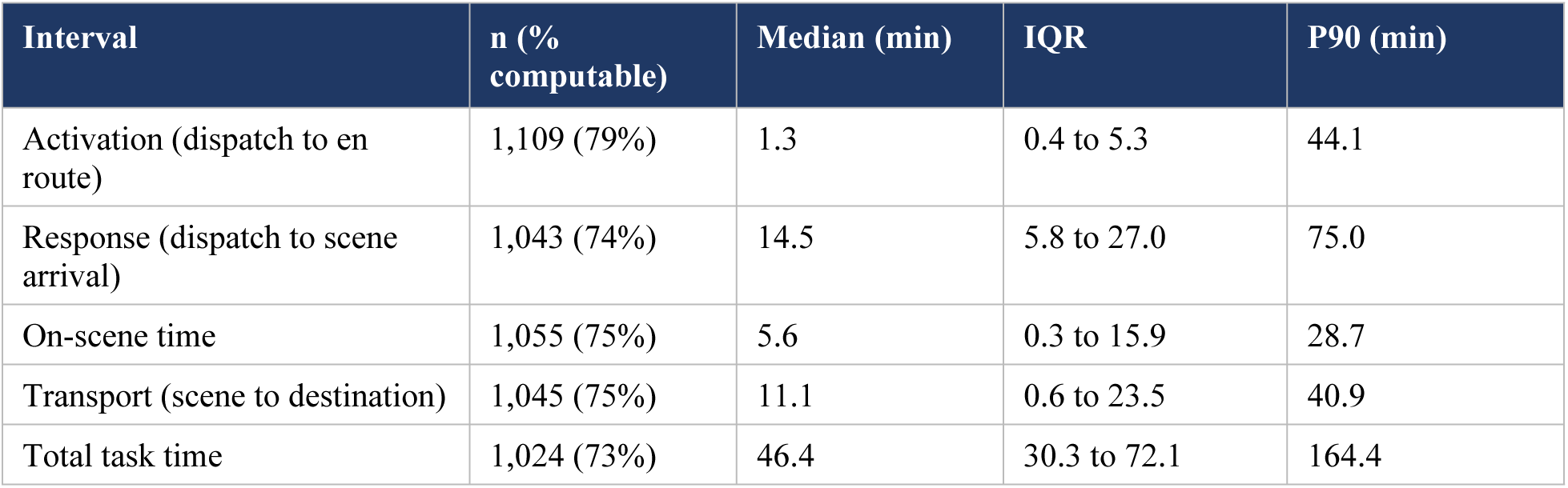
Prehospital response interval distributions, HEARTS first operational phase, Addis Ababa (trips with computable timestamps).

| Interval | n (% computable) | Median (min) | IQR | P90 (min) |
| --- | --- | --- | --- | --- |
| Activation (dispatch to en route) | 1,109 (79%) | 1.3 | 0.4 to 5.3 | 44.1 |
| Response (dispatch to scene arrival) | 1,043 (74%) | 14.5 | 5.8 to 27.0 | 75.0 |
| On-scene time | 1,055 (75%) | 5.6 | 0.3 to 15.9 | 28.7 |
| Transport (scene to destination) | 1,045 (75%) | 11.1 | 0.6 to 23.5 | 40.9 |
| Total task time | 1,024 (73%) | 46.4 | 30.3 to 72.1 | 164.4 |

Provider-level response intervals are reported in Table 1. The city-administrative public provider had a median response time of 16.1 minutes (n=804 computable). The hospital-based provider had a median response time of 8.6 minutes (n=202). The central coordination dispatch category had a median response time of 1.5 minutes (n=34). Prior to HEARTS, no unified, cross-provider prehospital response-time data existed for Addis Ababa.

### Documentation completeness

Timestamp field completeness ranged from 73% to 79% across interval types. Clinical field completeness was substantially lower: consciousness was recorded in 6.2% of trips, AVPU response in 2.0%, breathing rate in 3.9%, pulse rate in 2.3%, blood pressure in 1.9%, and primary management given in 0.0% of trips. Age was recorded in 68.7% of trips. Sex was recorded in 0.9% of trips.

### Implementation challenges and responses

Key implementation challenges encountered during the development and first operational phases of HEARTS are documented below, along with the responses implemented by the EAAN team.

### Challenge 1: Provider fragmentation and siloed operations

Five distinct provider types across Addis Ababa operated with independent dispatch systems, separate emergency access numbers, and no cross-provider data sharing infrastructure. No single regulatory or operational framework connected them. Each provider had its own call-centre protocols and patient care documentation practices, developed independently over years of isolated operation.

### Challenge 2: Resistance to digital transformation and data sharing

Private dedicated ambulance providers resisted platform registration and call logging. Concerns centred on commercial confidentiality: sharing trip data and call volumes with a unified platform was perceived as a threat to competitive positioning. Resistance was also observed among some public-sector dispatch personnel who were unfamiliar with digital workflows and uncertain about the accountability implications of recorded performance data.

### Challenge 3: Limited call-centre infrastructure

Several provider types, especially among the private providers, lacked the physical and technical infrastructure to support digital dispatch operations, including reliable computers, stable internet connectivity, and structured call-centre workflows. Some sub-city AAFDRMC stations had intermittent power supply and limited bandwidth, creating functional barriers to consistent platform use.

### Challenge 4: Inconsistent emergency care documentation

Paper-based patient care records varied substantially in structure and content across provider types. No standardised minimum dataset existed for prehospital clinical documentation, resulting in the low clinical field completeness rates observed in the HEARTS data. The absence of a national prehospital data standard complicated efforts to build a unified digital documentation workflow.

### Challenge 5: Connectivity constraints

Mobile network coverage gaps in certain sub-cities, particularly in peri-urban areas, led to intermittent GPS tracking failures and delayed data synchronisation between ambulance units and the central HEARTS platform. Offline data entry modes were required for some operational contexts.

### Challenge 6: Workforce capacity for system development and operation

Ethiopia had limited prior experience in developing and operating integrated multi-provider digital dispatch platforms. Building internal technical capacity for software customisation, server management, and user training required sustained investment and was constrained by the limited pool of locally experienced personnel in this domain.

### Challenge 7: Absence of a unified policy and financing framework

No existing policy framework in Ethiopia mandated participation in a shared prehospital coordination platform, and no dedicated financing mechanism existed for multi-provider coordination infrastructure. HEARTS was developed without a formal interoperability mandate, relying on voluntary participation and goodwill agreements.

### Responses to challenges

The EAAN team applied the following responses to these challenges during the study period:

- Telecommunications partnership: A formal partnership was established with a major Ethiopian telecommunications provider to negotiate preferential data plans for ambulance units and to address connectivity constraints in lower-coverage sub-cities.
- Internal capacity building: An internal technical team was trained and developed for platform development, management, customisation, and first-line user support, reducing dependence on external vendors and building sustainable domestic expertise.
- Incentivisation mechanisms: Performance-based visibility mechanisms were introduced for participating providers, including sub-city-level dashboard rankings and quarterly performance reports, to encourage data entry and demonstrate the value of participation to provider managers.
- Bilateral engagement with private providers: Structured bilateral negotiations with private dedicated ambulance providers addressed commercial confidentiality concerns through tiered data governance arrangements, allowing aggregate performance reporting without disclosure of commercially sensitive call-volume data.
- Funding diversification: Applications were submitted to multiple domestic and international funding sources to support platform development and operational costs. Local sustainability mechanisms, including potential cost-sharing arrangements with receiving hospitals and municipal health budgets, are under development.
- Stakeholder-driven Delphi process: A modified Delphi consensus process was initiated with Ethiopian prehospital experts to develop a nationally endorsed minimum dataset for prehospital documentation, addressing the absence of data standards.

Despite these responses, several challenges remained only partially resolved at the close of the study period, as reflected in the completeness of documentation and private provider participation data reported above.

## Discussion

This paper reports the implementation of HEARTS and EAAN in Addis Ababa, describes the challenges encountered, and presents early operational data as a contribution towards operationalising WHA 76.2’s integrated emergency care mandate. Our experience parallels and extends lessons from multicentre digital quality registries in resource-constrained settings (24,25), and from digital coordination platforms in other African cities(11,12,26). We draw five lessons that we believe are transferable to African cities facing similar provider fragmentation.

### Lesson 1: Governance before technology, and trust before governance

The central implementation challenge in PRECOS-1 was institutional, not technical. Providers accustomed to independent operation required sustained engagement before they would share a platform. Private dedicated ambulance providers resisted data registration due to well-founded commercial concerns; public-sector dispatchers expressed uncertainty about accountability implications of recorded performance data; and the absence of a unified policy framework meant that EAAN had no regulatory authority to mandate participation, relying entirely on voluntary engagement. This mirrors findings from global qualitative analyses of emergency care, which identify institutional fragmentation and lack of coordinating authority as primary barriers to EMS development in LMICs (27). The resolution in PRECOS-1 was a combination of bilateral negotiation, tiered data governance arrangements that protected provider confidentiality, and performance-based incentives that made participation visibly valuable. As demonstrated in the NaPQIN peri-operative registry (24), a clinician- and institution-led collaborative approach that respects provider autonomy is more sustainable than an externally imposed technical mandate. African cities or other LMICs planning similar reforms should anticipate that governance negotiations will take longer than technical deployment.

### Lesson 2: Infrastructure constraints are real and require targeted solutions

Limited call-centre infrastructure, connectivity gaps in peri-urban sub-cities, and the absence of standardised documentation practices were not peripheral challenges but central ones, shaping which providers could participate fully and which could not. The low clinical documentation completeness (0.0 to 6.2% across clinical fields) directly reflects the absence of a national minimum prehospital dataset and the absence of trained clinical documentation personnel on ambulance crews. This is a pre-standard-state problem: HEARTS captures what providers currently document and cannot generate data fields that no crew is trained or mandated to complete. The WHO’s 2024 operational guidance for ambulance systems specifically mandates standardised clinical documentation as a quality requirement (5). Building local technical capacity, establishing telecommunications partnerships to ensure connectivity, and developing an Ethiopia-specific, internationally comparable minimum dataset through a concurrent Delphi process constitute the structural responses to these challenges. They are prerequisites for PRECOS-2’s clinical outcome analysis and for any future before-and-after comparison.

### Lesson 3: The coordination layer operationalises WHA 76.2 and generates the first measurable baseline

Prior to HEARTS, no cross-provider prehospital response time data existed for Addis Ababa. The median response time of 14.5 minutes (IQR 5.8 to 27.0; P90 75.0 minutes) constitutes the first evidence-based operational benchmark for prehospital response in this city. The P90 response time of 75.0 minutes is the most actionable single finding from PRECOS-1: it identifies a subset of substantially delayed responses that were previously invisible and now constitute a specific quality improvement target. This is consistent with the wide variance in LMIC response times documented in the systematic review literature (7). WHA 76.2 calls for strengthened communication, transportation, and referral linkages, precisely what HEARTS provides through unified dispatch, real-time fleet tracking, and a single public access point (4). The Global Prehospital Consortium’s Delphi study places communication and coordination at the highest priority tier for LMIC EMS development (14), and PRECOS-1 demonstrates that this infrastructure can be built within a two-year timeline. The differential median response times between the city-administrative public provider (16.1 minutes) and the hospital-based provider (8.6 minutes) reflect fundamentally different dispatch-to-departure pathways for different mission types, a distinction that only unified multi-provider data makes visible. The real-time dashboard, disaggregated by sub-city and provider, now enables dispatch coordinators to identify underperforming locations and redeploy resources, a function that was structurally impossible before the platform existed. Future PRECOS phases should explore AI-assisted dispatch forecasting, which a recent scoping review identified as the most commonly studied digital health application for LMIC prehospital systems(28).

### Lesson 4: Maternal and obstetric care is the highest-impact early use case, and community engagement is inseparable from impact

Maternal and obstetric presentations constituted 44.3% of all coordinated trips, the largest single clinical category. Ethiopia’s maternal mortality ratio remains among the highest globally (estimated at 412 per 100,000 live births in 2016 (29), with HSTP-II targeting 279 per 100,000 by 2025 (15). The three delays model identifies transport as the primary bottleneck in the second delay, that of reaching care, which coordinated ambulance dispatch directly addresses (30). Research from South Africa confirms that delays in prehospital response to pregnancy-associated haemorrhage increase maternal mortality, and that coordinated resource allocation can reduce these delays (31).

The patient account illustrates the demand-side dimension: his wife’s obstetric emergency was reached in under five minutes at midnight, and he was getting constant updates on the ambulance arrival. His subsequent advocacy to his community exemplifies the multiplier effect that a single positive experience can generate. Community awareness of emergency access numbers and confidence in system response are as important as supply-side coordination: each underpins the other. Future PRECOS phases will systematically evaluate changes in ambulance utilisation for obstetric emergencies following community outreach, providing the evidence base needed to justify sustained investment in community engagement alongside platform development.

### Lesson 5: Formal enrolment and operational integration are distinct stages, each requiring targeted intervention

Private dedicated ambulance providers contributed 0.4% of coordinated trips despite being enrolled in the governance framework. The NGO humanitarian provider’s extensive volunteer ambulance network, while incorporated in governance, had not yet completed technical integration by the close of the study period. This gap between enrolment and operational integration is a known pattern in multi-stakeholder health network implementation (22), and reflects the different incentive structures, workflow requirements, and data governance concerns of private and NGO providers compared to public services. Resolving these gaps for PRECOS-2 requires revised onboarding protocols, bilateral negotiations on data governance, and outcome-sharing arrangements that demonstrate tangible benefit to participating providers. The NGO humanitarian provider’s activation will substantially expand the platform’s geographic reach into communities currently served only by volunteer networks.

### A programmatic research platform: from PRECOS-1 to longitudinal evidence

PRECOS-1 establishes the foundational measurement infrastructure for a research programme on prehospital coordination in Ethiopia. PRECOS-2 will conduct a before-and-after comparison of response intervals using historical paper-based dispatch records from AAFDRMC sub-city stations, generating the first empirical estimate of the effect of HEARTS on response time. PRECOS-3 will conduct a prospective community utilisation survey to examine changes in ambulance awareness and call rates, with a particular focus on obstetric emergencies. Infrastructure: extend the HEARTS platform to remote and rural regions of Ethiopia, where geographic isolation and infrastructure constraints create a prehospital challenge distinct from the urban Addis Ababa context; findings from this expansion will test whether the coordination-first governance model transfers across the urban-rural continuum. The concurrent Ethiopia-focused Delphi process will strengthen the variable framework for subsequent phases. As the Ethiopian evidence base matures, the programme continues to seek South-South collaborations with EMS networks in Kenya, Rwanda, and other African settings for comparative analyses of fragmentation patterns and coordination solutions and ultimately help establish the continental-level prehospital ambulance alliance research network.

Taken together, the EAAN/HEARTS model addresses the three barriers the African Federation for Emergency Medicine identified as holding back prehospital progress across the continent: perceived cost, absence of an agreed-upon system structure, and poor advocacy (21). Coordination-first is cheaper than fleet expansion; EAAN provides the governance framework, and a federated multi-provider network is itself an advocacy structure for prehospital investment.

### Limitations

There are some limitations that constrain the interpretation of PRECOS-1 findings. The study is an early implementation account without a control group or pre-implementation performance comparator, and no causal inference about outcomes can be drawn. Participation by private providers and NGO humanitarian providers was minimal, limiting current generalisability to the full multi-provider landscape. Clinical documentation completeness was very low across most fields, precluding any clinical outcome analysis in this phase. Additionally, no patient-reported outcomes were collected during this phase; the patient experience reported in the Results represents a single consented account rather than a systematic assessment of patient experience. Response intervals were computable for 74% of trips; if missingness is non-random across provider or acuity type, reported medians may not represent all trips. Finally, PRECOS-1 is a single-city study, reporting only from Addis Ababa; the HEARTS platform is expanding to remote and rural regions of Ethiopia, and findings from those settings will be reported in subsequent PRECOS studies.

## Conclusions

In resource-constrained settings where fragmentation of prehospital providers results in unmeasured and uncoordinated emergency care, digital coordination platforms linked to multi-stakeholder governance can render prehospital performance measurable, actionable, and continuously improvable. PRECOS-1 demonstrates that a coordination-first approach to operationalising WHA 76.2’s integrated emergency care mandate is feasible within a two-year implementation timeline, even in a highly fragmented setting with significant infrastructure constraints, provider resistance, and the absence of a unified policy framework. The first unified prehospital response baseline for Addis Ababa, a maternal and obstetric caseload representing 44% of all coordinated trips, and a P90 response time of 75 minutes, identifying where the system currently fails: these findings establish the measurement foundation against which all future improvement in PRECOS-2 and beyond will be judged.

The implementation challenges documented in PRECOS-1, including provider resistance, connectivity gaps, the absence of data standards, and financing uncertainty, are not unique to Ethiopia. They are the shared landscape of prehospital coordination reform across Africa. The responses developed by the EAAN team, including telecommunications partnerships, bilateral governance agreements, performance incentives, and internal capacity building, represent a practical toolkit transferable to other settings. EAAN’s model, combining multi-stakeholder governance, shared digital dispatch, real-time performance dashboards, a community-facing mobile application, and a single public access point, is not Ethiopia-specific. It is a replicable, evidence-based response to a continent-wide problem, and PRECOS provides the research and quality improvement infrastructure to test, refine, and scale it.

## Declarations

### Author contributions

Conceptualisation: FK. Methodology: PKD, KRI, BK, EDA, OAN, EBT, FK. Formal analysis: FK. Software: BK. Data curation: PKD, BK. Resources: BK, ET, TK, EBT. Investigation: PKD, KRI, AA, ET, EDA, HM, NZ, OAN, RAA, TK, KD, YB EBT. Project administration: PKD, BK, EDA, KD, FK. Supervision: KRI, EBT, FK. Writing original draft: FK. Writing review and editing: all authors. All members of the writing committee and the EAAN Collaborative read and approved the final version of the manuscript submitted for publication.

### Ethics approval

AHRI/ALERT Ethics Review Committee, protocol PO-039/25, approval period 18 July 2025 to 17 July 2026. Patient-identifiable fields were excluded from all analyses. The patient account is reproduced with the full written consent of the patient and family.

### Data availability

De-identified operational data will be made available upon reasonable request to the corresponding author, subject to the EAAN data governance framework and ethics approval conditions.

## Acknowledgements

The authors thank all members of the Ethiopian Ambulance Alliance Network (EAAN) Collaborative for their contributions to this work. A full list of Collaborative members is provided in the supplementary material.

## EAAN Collaborative membership

(Provide full names and surnames of all Collaborative members for the supplementary file, required for PubMed indexing following the NaPQIN model.)

## AI use declaration

AI-assisted tools were used to support data analysis and word/grammar editing. All content, analyses, interpretations, and conclusions represent the authors’ own intellectual work, reviewed and approved by all authors. AI tools were not used to generate or verify references.

